# Problem drinking before and during the COVID-19 crisis in US and UK adults: Evidence from two population-based longitudinal studies

**DOI:** 10.1101/2020.06.25.20139022

**Authors:** Michael Daly, Eric Robinson

## Abstract

**Background:** The impact of the COVID-19 crisis on potentially harmful alcohol consumption is unclear.

**Aims:** To test whether the prevalence of problem drinking has changed from before to during the COVID-19 crisis in the US and UK.

**Design/Setting:** We examined nationally representative longitudinal data on how problem drinking has changed from pre-pandemic levels among adults in the US (N=7,327; Understanding America Study) and UK (N=12,594; UK Household Longitudinal Study).

**Methods:** In the US, we examined rates of consuming alcohol ≥ 4 times in the past week at baseline (March, 2020) and across four waves of follow-up (April-May, 2020). In the UK we assessed the prevalence of consuming alcohol ≥ 4 times per week and weekly heavy episodic drinking using the AUDIT-C at baseline (2017-2019) and during the COVID-19 lockdown (April, 2020). We also tested whether there were specific groups at greater risk of increased problem drinking during the pandemic.

**Results:** Among US adults, there was a statistically significant increase in the percentage of participants reporting drinking alcohol ≥ 4 times a week which rose significantly from 11.7% to 17.9% (53% increase, *p* < .001) as the COVID-19 crisis developed in the US. Among UK adults, the percentage of participants reporting drinking ≥ 4 times a week increased significantly from 14.2% to 23% (62% increase, *p* < .001) and heavy episodic drinking at least weekly increased significantly from 9.7% to 16.6% (71% increase, *p* < .001) when compared to pre-COVID-19 lockdown levels. Trends were similar across population demographics, although those aged under 50 years and higher income groups displayed the largest increases.

**Conclusions:** The COVID-19 crisis has been associated with substantial increases in problematic drinking in both US and UK adults.

## INTRODUCTION

The COVID-19 pandemic has resulted in governments introducing drastic measures to reduce viral transmission. Many governments have introduced ‘social lockdown’ orders, which have had severe effects on the economy and far reaching interpersonal consequences on working life, childcare, travel and social contact. Although social lockdown orders will have reduced the number of deaths caused by COVID-19, as of June 2020, in the UK alone there have been more than 30,000 deaths attributed to COVID-19 (1, 2). There is also emerging evidence on the indirect effects the COVID-19 pandemic has had on population health. For example, initial findings from both the UK and the US indicate that the COVID-19 pandemic is likely to have impacted mental health, with substantial increases in the prevalence of mental health problems and depression estimated from nationally representative studies (3, 4).

The extent to which alcohol use has changed as a result of COVID-19 crisis is unclear. There is already a considerable public health burden caused by problematic drinking (5) and alcohol misuse could increase risk of mortality from COVID-19 because of immune function related health effects (6). For these reasons, it is crucial to understand how patterns of problematic drinking have changed since the emergence of the COVID-19 crisis. In the US and UK, there have been mass closures of non-essential businesses, including pubs, bars and restaurants, which may have reduced the amount of alcohol that the population are drinking. However, this has also coincided with a sharp rise in alcohol sales in supermarkets (7). There are also concerns that COVID-19 social lockdown measures may result in a spike in alcohol misuse, particularly among groups that are already at risk for problematic drinking patterns (8, 9).

Prior research has shown that exposure to traumatic events such as Hurricane Katrina (10) and the 9/11 terrorist attacks (11) predicts alcohol misuse and drinking to alleviate distress and worry related to the event. Yet, research studies examining problematic drinking during the COVID-19 crisis are limited. In two non-representative cross-sectional studies relying on retrospective recall of alcohol drinking prior to the COVID-19 pandemic, approximately one quarter of Australian adults and one third of Chinese adults reported that their alcohol consumption had increased as a result of COVID-19 lockdown (12, 13). In a repeated cross-sectional survey, there was an increase in the prevalence of high-risk drinking of approximately 50% among 1700 UK adults (14) when comparing drinking before and after COVID-19 social lockdown. Although these studies are suggestive of changes in problematic drinking, findings may be explained by the use of retrospective recall and/or differences between participants sampled before vs. during the COVID-19 crisis. Therefore, there is a need for research that allows for examination of longitudinal changes in person-by-person problematic drinking behavior before and after the development of the COVID-19 pandemic.

In the present research we examine changes in problematic drinking among US and UK adults before and during the development of the COVID-19 pandemic. We make use of two longitudinal studies with well characterized sampling frames and sampling weights that provide a correction for selection probabilities and attrition bias enabling population inferences to be generated. We examined problematic drinking patterns among US adults by making use of data collected as part of the Understanding America Study. In this study, drinking behavior was reported on early in the COVID-19 pandemic and before social lockdown restrictions had been widely introduced in the US (March, 2020) vs. during lockdown restrictions (April, 2020) and after the easing of restrictions (May, 2020). We also examined drinking patterns among UK adults by making use of data collected as part of the UK Household Longitudinal study in 2017-2019 and again in April, 2020 one month after the introduction of UK-wide lockdown restrictions. To understand whether trends in problematic drinking were socially patterned, we also examined changes in problem drinking based on demographic sub-groups (age, sex, ethnicity, marital status, and income).

## METHODS

### Participants

This study used data from two nationally representative longitudinal studies: the Understanding America Study (UAS) and the UK Household Longitudinal Study (UKHLS or *Understanding Society*). The UAS is a probability-based internet panel where those without initial internet access are provided with tablet computers to ensure representativeness. The study began in 2014 and participants were recruited via address-based sampling from the US Postal Service Computerized Delivery Sequence file covering almost 100% of US households (15). In March, 2020 8,547 participants from the UAS were invited to take part in a COVID-19 Tracking Study and 7,420 agreed.

In this study we use data from 7,327 participants who provided 30,966 observations over five waves of data collection conducted fortnightly from early March to the end of May. The first wave of the survey was fielded from March 10^th^ to 31^st^ with 85% of participants completing the survey by March 19^th^ when California introduced the first stay-at-home order. Most US states followed suit enacting social lockdown measures in the two-week period that followed (16). A rapid increase in COVID-19 cases took place from March 19^th^ to April 1^st^ when the number of confirmed cases per day increased from approximately 5,000 to over 25,000 in the US (17).

Four subsequent survey waves have been conducted as part of the UAS COVID-19 Tracking Study over 14-day periods from April 1-14, April 15-28, April 29-May 12, and May 13-26. Each participant was assigned a day to complete their survey during each wave and 93.3% did so on their assigned day (18). In this study we include the remaining surveys that were not completed on the assigned day but were completed within two weeks of the assigned date. Sampling weights were applied in all analyses to adjust for non-response and generate nationally representative estimates. In the UAS survey-wave specific sampling weights are generated using an adaptive sampling algorithm described elsewhere (19). The weights account for unequal probabilities of selection into the UAS and ensure each wave of the study is aligned with the distribution of sociodemographic characteristics of the US population.

The UKHLS is a longitudinal study that collects extensive information annually on the health and economic circumstances of UK households. The sample combines a general population sample, ethnic minority boost samples and participants from the British Household Panel Study (BHPS) all recruited via stratified equal probability sampling of addresses from across the UK selected from the Postcode Address File. Fieldwork for each wave of the UKHLS takes over two years and survey waves partly overlap. In the current study, we draw on data from the latest (Wave 9: N =32,596) sweep of the UKHLS that ran from January 2017 to May 2019 and had a response rate of 67.9%.

Data from this wave was merged with the UKHLS COVID-19 study that ran from 24-30^th^ April one month after the introduction of a stay-at-home order in the UK on March 23^rd^. The survey was completed by 46% of Wave 9 participants (N = 14,985) (20). The number of completed COVID-19 interviews with survey weights available to provide nationally representative estimates was 13,704 and of this group 1,110 were missing data on one or more of the study outcomes or demographic variables giving a final sample size of 12,594. A small portion of income assessments were missing (N =227; 1.8%) and were replaced with a missing data dummy variable. Participant responses were reweighted using inverse probability weights developed using the rich demographic, health, and economic variables available in the representative Wave 9 wave of the UKHLS. This provided an adjustment for both unequal selection probabilities and non-random non-response to the COVID-19 survey among those who completed the Wave 9 survey (21).

### Measures

#### Problem drinking

In each wave of the UAS COVID-19 Tracking study participants were asked “Out of the past 7 days, what is your best estimate of the number of days that you did each of the following activities?” and were asked to complete the number of days they “Consumed alcohol” alongside other health behaviors. To identify potentially problematic drinking and enable comparisons with the UKHLS we dichotomized responses to this question into those who drank more or less frequently than 4 times in the past week.

In the UKHLS participants completed the AUDIT-C (22) in 2017-2019 and again in April, 2020. In 2017-2019 participants were asked “Thinking about the past 12 months, how often do you have a drink containing alcohol?” and responded on a four-point scale (1=Never, 2=2-4 times per month, 3=2-3 times per week, 4=4+ times per week). In order to capture drinking levels during the pandemic lockdown in the COVID-19 study the reference period for this question was changed from “past 12 months” to “past 4 weeks” and response scales were as follows: 1=Never, 2=Once, 3=2-4 times in total, 4=2-3 times per week, 5=4-6 times per week, and 6=Daily. In both waves, we characterized problematic drinking as consuming alcohol 4 or more times per week.

The AUDIT-C also includes a question on the frequency of heavy episodic alcohol use, defined for women/men as drinking 6/8 or more units on a single occasion: “How often have you had 6 or more units (if sex =female) / 8 or more units (if sex =male), on a single occasion in the last year?” Participants were instructed that “By a unit we mean ½ pint of beer, a glass of wine or a single measure of spirit or liquer.” In 2017-2019 the reference period was “the past year” and response options were: 1=Never, 2=Less than monthly, 3=Monthly, 4=Weekly, 5=Daily or almost daily. Once again in April, 2020 the reference period referred to was adapted from “the past year” to “the past 4 weeks” and heavy episodic drinking was assessed with the response options: 1=Never, 2=Once, 3=Weekly, 4=Daily or almost daily. In both 2017-2019 and April, 2020 those indicating that they drank heavily on a ‘Weekly’ or ‘Daily or almost daily’ basis were classified as engaging in problem drinking.

#### Covariates

In both the UAS and UKHLS participants reported their age, sex, ethnicity (grouped into white, non-white due to a low proportion of Black, Asian, and minority participants in the UKHLS), marital status (married, not married) and household income levels. Participants were grouped into four approximately even sized age groups (18-34, 35-49, 50-64, 65+) and three household income groups (UAS: ≤$40,00, $40,000– $100,000 ≥$100,000 gross per annum; UKHLS: ≤£2,500, £2,500–£4,000, ≥£4,000 net income per month).

### Statistical analysis

Our analyses of both datasets incorporated survey sampling weights to produce representative estimates. First, we outlined the descriptive trends in problem drinking levels across survey waves for all participants and population subgroups (i.e. age, sex, race/ethnicity, marital status, and income groupings). Next, both studies were examined separately using logistic regression analysis to test for the presence of differences in the prevalence of frequent drinking and heavy episodic drinking between the first survey wave which was treated as a baseline (UAS: March, 2020; UKHLS: 2017-2019) and subsequent survey waves which were treated as follow-up assessments (UAS: four waves across April and May, 2020; UKHLS: April, 2020 wave).

To do this, we first estimated the predicted probability of each problem drinking outcome at each survey wave in logistic regression models that adjusted for differences in participant age, sex, ethnicity (white, non-white), marital status (married, not married), and household income tertiles. The Stata ‘margins’ command was then used to estimate percentage-point changes in the binary outcomes of interest from the first survey wave / baseline to subsequent follow-up waves while adjusting for the distribution of covariates (23). Standard errors were clustered by the individual participant identifier to account for repeated observations across waves in both studies.

Next, we sought to examine patterns of change in levels of problem drinking across population subgroups. Interactions between the survey wave variable and each demographic variable were introduced into the model and estimated simultaneously. The margins command was then used to estimate discrete change in levels of problem drinking from the first survey wave / baseline to subsequent follow-up assessments for each subgroup. Finally, we used the Stata lincom command to test for the presence of systematic differences in patterns of change in problem drinking across waves between population subgroups (e.g. to test if the magnitude of an increase in heavy episodic drinking from 2017-2019 to April, 2020 in the UKHLS was larger in high income participants compared to low income participants).

## RESULTS

### Understanding America Study

The sample for our analyses included 7,327 participants assessed over five waves (Ns from 5,395–6,819). Participants were aged 48.5 (95% CI[48-49.1]) and 53.6% were female and 66% white, as shown in Table 1. The characteristics of participants in each survey wave are outlined in Table S1 which shows that the weighted sample composition was very similar in each wave. The prevalence of drinking four or more times in the past week was 11.7% in the first survey wave conducted in March, 2020 and increased to 17.8% in early April and remained elevated at 16.5% by late May, 2020 (see Table 1). The increase in frequent drinking was largest in magnitude amongst those aged under 50 years, whites, those who were not married, and those living in households earning $40,000 or more per year.

**Table 1.**
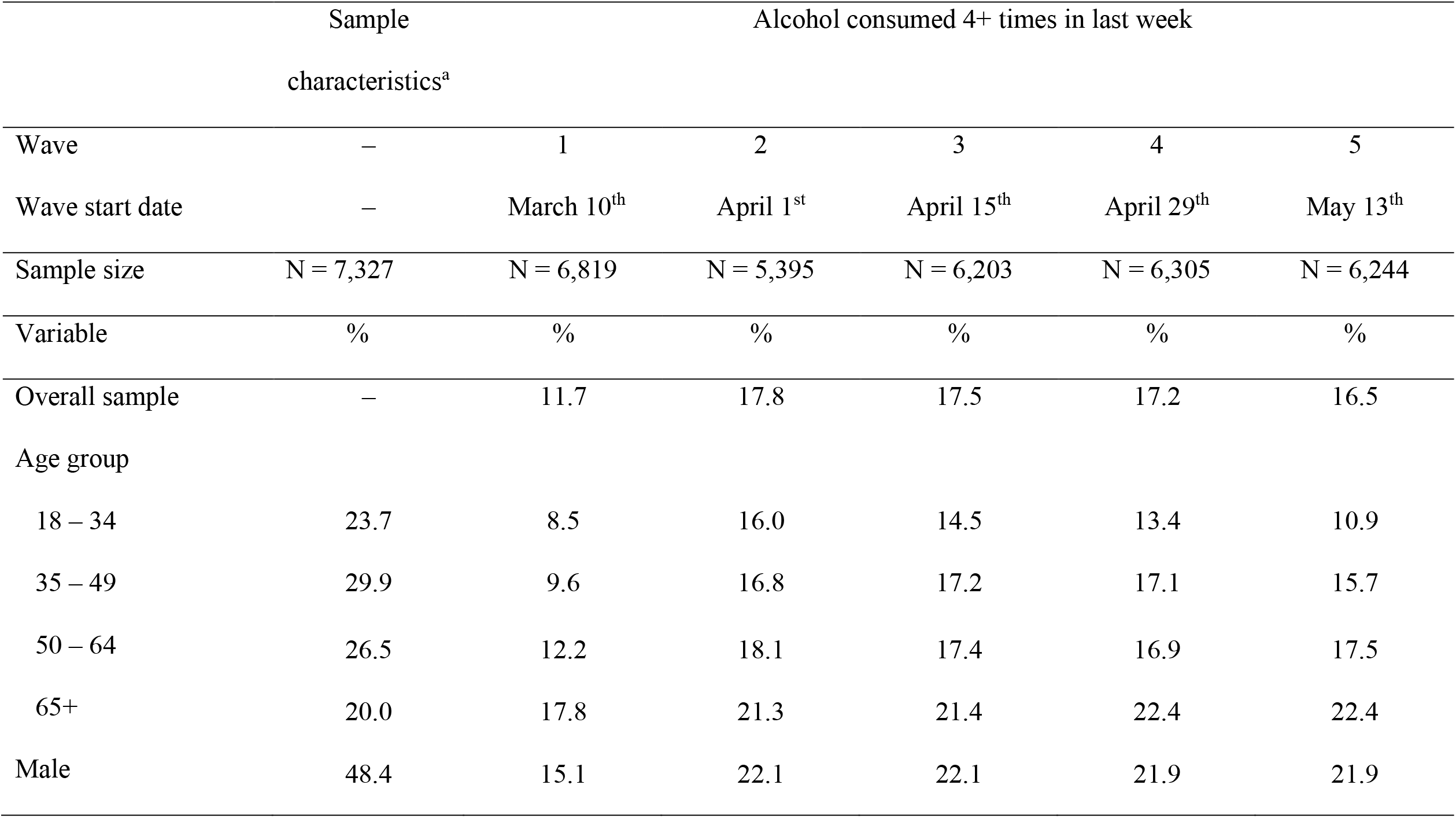
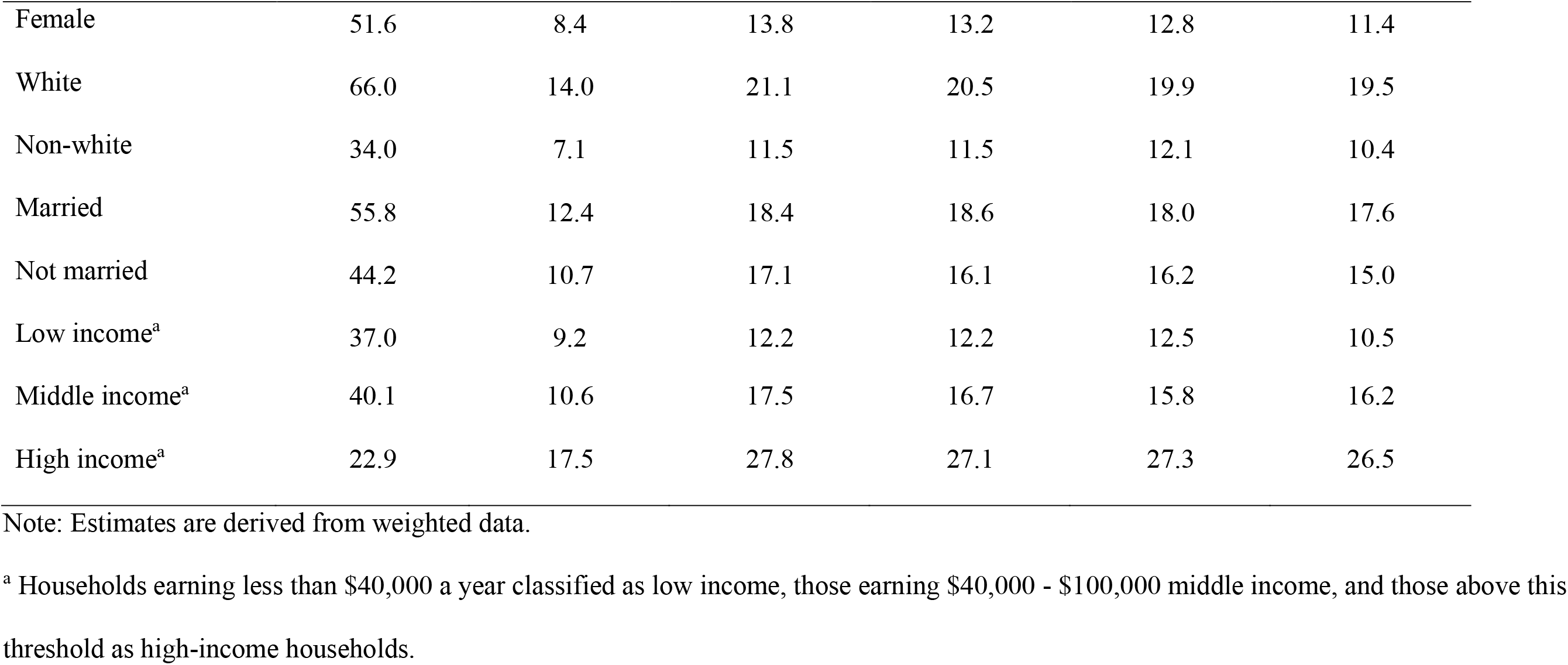
Sample characteristics and the prevalence of drinking alcohol 4+ times in the past week before (March, 2020) and during the COVID-19 pandemic (April, 2020) in five waves of the Understanding America Study (UAS) (N = 7,327; Obs. = 30,966).

|  | Sample | Alcohol consumed 4+ times in last week |  |  |  |  |
| --- | --- | --- | --- | --- | --- | --- |
|  | characteristics<sup>a</sup> |  |  |  |  |  |
| Wave | – | 1 | 2 | 3 | 4 | 5 |
| Wave start date | – | March 10<sup>th</sup> | April 1<sup>st</sup> | April 15<sup>th</sup> | April 29<sup>th</sup> | May 13<sup>th</sup> |
| Sample size | N = 7,327 | N = 6,819 | N = 5,395 | N = 6,203 | N = 6,305 | N = 6,244 |
| Variable | % | % | % | % | % | % |
| Overall sample | – | 11.7 | 17.8 | 17.5 | 17.2 | 16.5 |
| Age group |  |  |  |  |  |  |
| 18 – 34 | 23.7 | 8.5 | 16.0 | 14.5 | 13.4 | 10.9 |
| 35 – 49 | 29.9 | 9.6 | 16.8 | 17.2 | 17.1 | 15.7 |
| 50 – 64 | 26.5 | 12.2 | 18.1 | 17.4 | 16.9 | 17.5 |
| 65+ | 20.0 | 17.8 | 21.3 | 21.4 | 22.4 | 22.4 |
| Male | 48.4 | 15.1 | 22.1 | 22.1 | 21.9 | 21.9 |
| Female | 51.6 | 8.4 | 13.8 | 13.2 | 12.8 | 11.4 |
| White | 66.0 | 14.0 | 21.1 | 20.5 | 19.9 | 19.5 |
| Non-white | 34.0 | 7.1 | 11.5 | 11.5 | 12.1 | 10.4 |
| Married | 55.8 | 12.4 | 18.4 | 18.6 | 18.0 | 17.6 |
| Not married | 44.2 | 10.7 | 17.1 | 16.1 | 16.2 | 15.0 |
| Low income<sup>a</sup> | 37.0 | 9.2 | 12.2 | 12.2 | 12.5 | 10.5 |
| Middle income<sup>a</sup> | 40.1 | 10.6 | 17.5 | 16.7 | 15.8 | 16.2 |
| High income<sup>a</sup> | 22.9 | 17.5 | 27.8 | 27.1 | 27.3 | 26.5 |

Note: Estimates are derived from weighted data.
<sup>a</sup> Households earning less than \$40,000 a year classified as low income, those earning \$40,000 - \$100,000 middle income, and those above this threshold as high-income households.

### Regression models

In an adjusted model, the predicted probability of drinking four or more times in the past week increased from 11.7% (95% CI[10.7%-12.6%]) in March to 17.9% (95% CI[16.6%-19.2%]) in early April, 2020 a statistically significant increase of 6.2% (95% CI[5.0%-7.5%]) (*p* <.001), as shown in Table 2. We also examined problem drinking among participants who completed their baseline survey before lockdown measures were enacted (completed assessments on 10^th^-19^th^ March). We compared the prevalence of drinking ≥4 times per week in this group with the high frequency drinking levels of the same group of participants as averaged across assessment waves from April 15^th^-May 13^th^. As can be seen in Table S2 the prevalence of frequent drinking in this group (N=5,430) increased from 11.6% (95% CI[10.5-12.7]) to 17.5% (95% CI[16.2-18.6]) from March 10^th^-19^th^ to April/May, a statistically significant increase of 5.9% (*p* <.001). Subgroup differences also aligned with those identified in our main analyses. This analysis indicated that the inclusion of participants whose baseline drinking was assessed after a small number of states had introduced lockdown measures was unlikely to affect the trends identified.

**Table 2.**
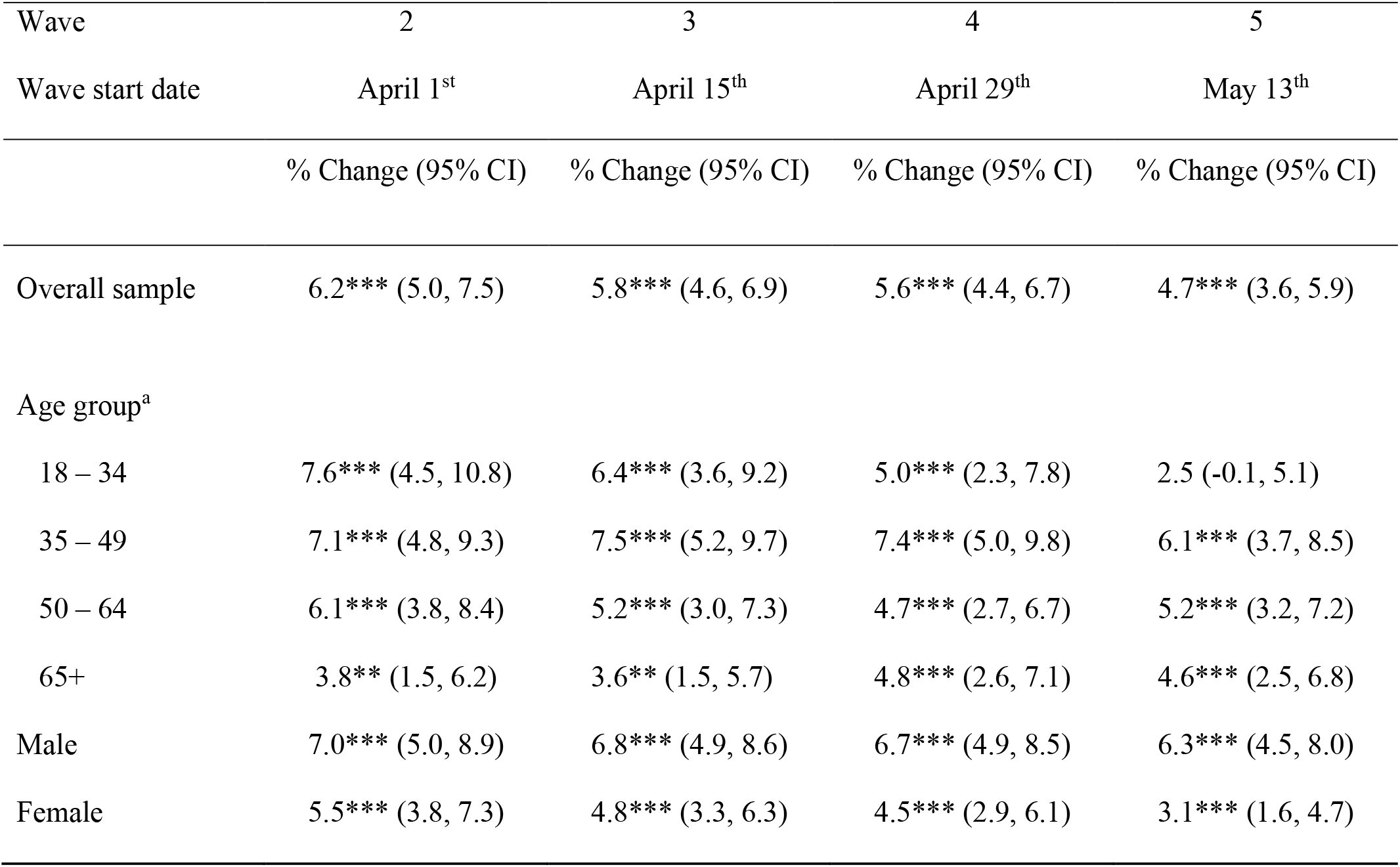
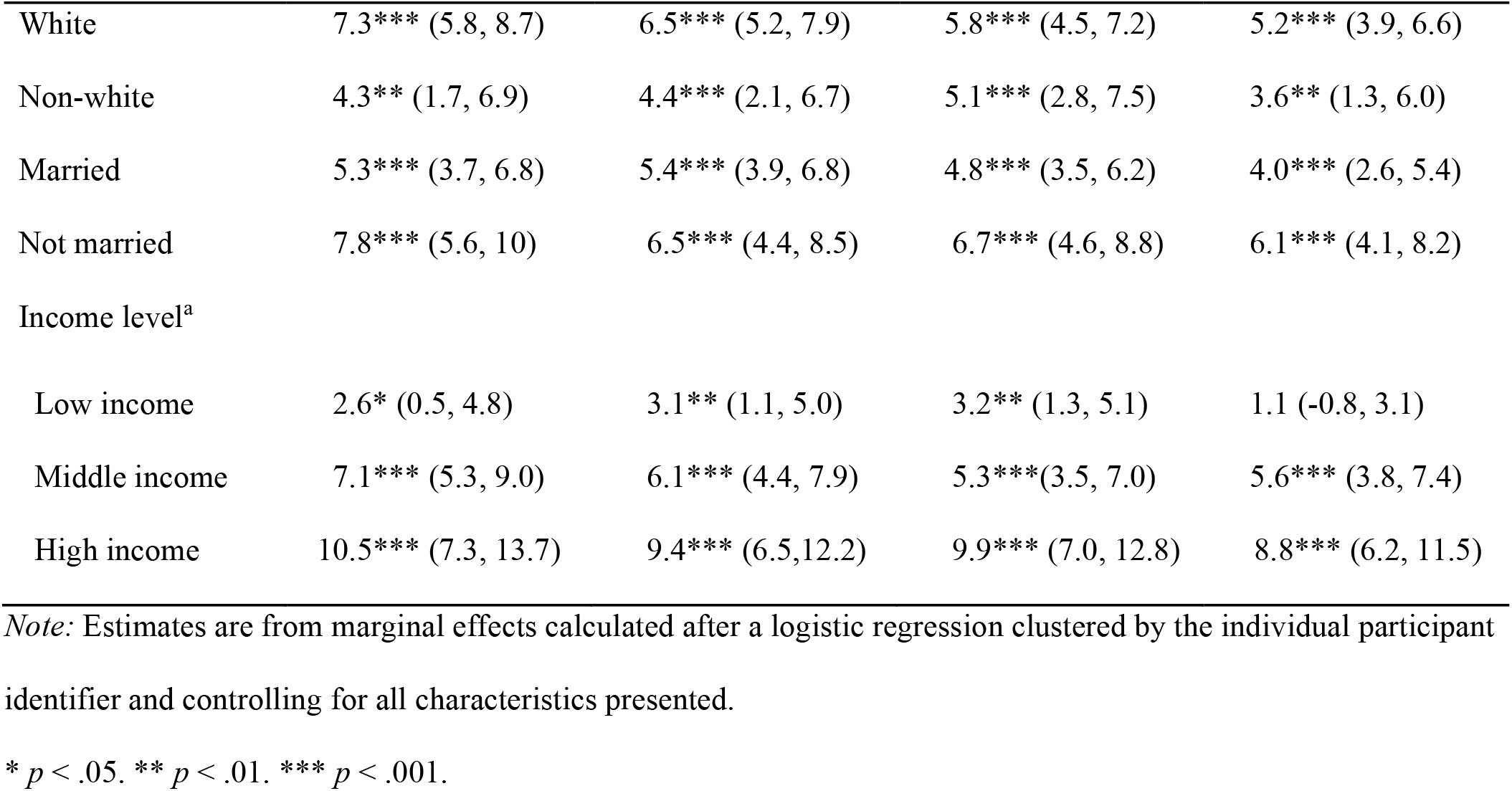
Regression estimates of percentage point changes in drinking 4+ times in the past week from March, 2020 to April-May 2020 in the Understanding America Study (UAS) (N = 7,327; Obs. = 30,966).

| Wave | 2 | 3 | 4 | 5 |
| --- | --- | --- | --- | --- |
| Wave start date | April 1<sup>st</sup> | April 15<sup>th</sup> | April 29<sup>th</sup> | May 13<sup>th</sup> |
|  | % Change (95% CI) | % Change (95% CI) | % Change (95% CI) | % Change (95% CI) |
| Overall sample | 6.2*** (5.0, 7.5) | 5.8*** (4.6, 6.9) | 5.6*** (4.4, 6.7) | 4.7*** (3.6, 5.9) |
| Age group<sup>a</sup> |  |  |  |  |
| 18 – 34 | 7.6*** (4.5, 10.8) | 6.4*** (3.6, 9.2) | 5.0*** (2.3, 7.8) | 2.5 (-0.1, 5.1) |
| 35 – 49 | 7.1*** (4.8, 9.3) | 7.5*** (5.2, 9.7) | 7.4*** (5.0, 9.8) | 6.1*** (3.7, 8.5) |
| 50 – 64 | 6.1*** (3.8, 8.4) | 5.2*** (3.0, 7.3) | 4.7*** (2.7, 6.7) | 5.2*** (3.2, 7.2) |
| 65+ | 3.8** (1.5, 6.2) | 3.6** (1.5, 5.7) | 4.8*** (2.6, 7.1) | 4.6*** (2.5, 6.8) |
| Male | 7.0*** (5.0, 8.9) | 6.8*** (4.9, 8.6) | 6.7*** (4.9, 8.5) | 6.3*** (4.5, 8.0) |
| Female | 5.5*** (3.8, 7.3) | 4.8*** (3.3, 6.3) | 4.5*** (2.9, 6.1) | 3.1*** (1.6, 4.7) |
| White | 7.3*** (5.8, 8.7) | 6.5*** (5.2, 7.9) | 5.8*** (4.5, 7.2) | 5.2*** (3.9, 6.6) |
| Non-white | 4.3** (1.7, 6.9) | 4.4*** (2.1, 6.7) | 5.1*** (2.8, 7.5) | 3.6** (1.3, 6.0) |
| Married | 5.3*** (3.7, 6.8) | 5.4*** (3.9, 6.8) | 4.8*** (3.5, 6.2) | 4.0*** (2.6, 5.4) |
| Not married | 7.8*** (5.6, 10) | 6.5*** (4.4, 8.5) | 6.7*** (4.6, 8.8) | 6.1*** (4.1, 8.2) |
| Income level<sup>a</sup> |  |  |  |  |
| Low income | 2.6* (0.5, 4.8) | 3.1** (1.1, 5.0) | 3.2** (1.3, 5.1) | 1.1 (-0.8, 3.1) |
| Middle income | 7.1*** (5.3, 9.0) | 6.1*** (4.4, 7.9) | 5.3*** (3.5, 7.0) | 5.6*** (3.8, 7.4) |
| High income | 10.5*** (7.3, 13.7) | 9.4*** (6.5, 12.2) | 9.9*** (7.0, 12.8) | 8.8*** (6.2, 11.5) |

*Note:* Estimates are from marginal effects calculated after a logistic regression clustered by the individual participant identifier and controlling for all characteristics presented.
\* $p < .05$ . \*\* $p < .01$ . \*\*\* $p < .001$ .

Statistically significant increases in the probability of frequent drinking from baseline to early April assessments were identified for all population subgroups examined. In line with descriptive trends, the statistically significant increases observed were largest among those aged under 50 (18-34: 7.6%, 35-49: 7.1%), whites (7.3%), those who were not married (7.8%) and those from households earning $40,000 or more per year (middle income: 7.1%, high income: 10.5%), as shown in Table 2. The increase in frequent drinking from March levels persisted throughout April and May in all subgroups except for those aged 18-34 and from households earning less than $40,000 per year which did not differ from baseline in late May (see Table 2).

Next, we tested whether the changes in levels of drinking four or more times per week from March to subsequent survey waves differed significantly between population subgroups. This analysis showed that those with high household incomes (≥$100,000) reported a 7.9% (95% CI[3.9%-11.9%], *p* <.001) greater increase in frequent drinking from March to early April than those on low incomes, an increase that persisted across all subsequent survey waves, as shown in Table 3. Similarly, those on middle incomes showed a rise in frequent drinking in early April that was significantly larger (by 4.5%, 95% CI[1.7%-7.4%]) than the increase experienced by those on low incomes. Among participants aged 35-49 the prevalence of drinking four or more times per week increased by 3.9% more (95% CI[0.8%-7%]) from March to late April, 2020 compared to those aged 65+.

**Table 3.** Regression estimates of subgroup differences in percentage point changes in drinking 4+ times in the past week from March, 2020 to April-May 2020 in the Understanding America Study (UAS) (N = 7,327; Obs. = 30,966).

| Wave | 2 | 3 | 4 | 5 |
| --- | --- | --- | --- | --- |
| Wave start date | April 1<sup>st</sup> | April 15<sup>th</sup> | April 29<sup>th</sup> | May 13<sup>th</sup> |
|  | % Change (95% CI) | % Change (95% CI) | % Change (95% CI) | % Change (95% CI) |
| Age group (comparison<br>group is 65+ years) |  |  |  |  |
| 18 – 34 | 3.8 (-0.2, 7.8) | 2.8 (-0.7, 6.3) | 0.2 (-3.3, 3.8) | -2.1 (-5.5, 1.2) |
| 35 – 49 | 3.2 (0.0, 6.5) | 3.9* (0.8, 7.0) | 2.6 (-0.7, 5.9) | 1.4 (-1.7, 4.6) |
| 50 – 64 | 2.2 (-0.9, 5.4) | 1.6 (-1.3, 4.5) | -0.1 (-3, 2.8) | 0.6 (-2.3, 3.5) |
| Male vs. female | 1.4 (-1.2, 4.1) | 1.9 (-0.5, 4.4) | 2.2 (-0.2, 4.6) | 3.1* (0.7, 5.5) |
| White vs. non-white | 3.0 (0.0, 5.9) | 2.2 (-0.5, 4.9) | 0.7 (-2.0, 3.5) | 1.6 (-1.1, 4.3) |
| Married vs. not married | -2.5 (-5.3, 0.3) | -1.1 (-3.7, 1.6) | -1.9 (-4.5, 0.7) | -2.2 (-4.8, 4.4) |
| Income level<sup>a</sup> |  |  |  |  |
| Middle vs. low | 4.5** (1.7, 7.4) | 3.1* (0.3, 5.8) | 2.1 (-0.5, 4.7) | 4.5** (1.8, 7.2) |
| High vs. low | 7.9***(3.9, 11.9) | 6.3** (2.7, 10) | 6.7***(3.1, 10.3) | 7.7***(4.3, 11.1) |

*Note:* Estimates are from marginal effects calculated after a logistic regression clustered by the individual participant identifier and controlling for all characteristics presented. Coefficients indicate the difference in the percentage point increase in drinking problems across survey ways between the groups examined. \* $p < .05$ . \*\* $p < .01$ . \*\*\* $p < .001$ .

### UK Household Longitudinal Study

Participants (N =12,594) were aged 51.3 (95% CI[50.9-51.7]), were 53.7% female, and 93.2% white, and 54.4% were married, as shown in Table 4. The prevalence of drinking 4+ times per week was 14.2% in 2017-2019 and rose to 23% in April, 2020. The prevalence of heavy drinking at least once a week rose from 9.7% in 2017-2019 to 16.6% in April, 2020. Changes in frequent drinking and heavy episodic drinking were largest in magnitude amongst those aged 35-49, females, whites, and those on middle or high incomes (see Table 4).

**Table 4.** Sample characteristics and the prevalence of problem drinking in the 2017-2019 and April, 2020 Waves of the UK Household Longitudinal Study (UKHLS; N = 12,594).

| Variable | Sample | Alcohol consumed |  | Heavy episodic |  |
| --- | --- | --- | --- | --- | --- |
|  | characteristics | 4+ times per week |  | drinking at least weekly |  |
|  |  | 2017-2019 | April, 2020 | 2017-2019 | April, 2020 |
|  | % | % | % | % | % |
| Overall sample | — | 14.2 | 23.0 | 9.7 | 16.6 |
| Age group<sup>a</sup> |  |  |  |  |  |
| 18 – 34 | 20.4 | 4.0 | 13.2 | 8.9 | 16.2 |
| 35 – 49 | 23.9 | 10.3 | 22.2 | 10.2 | 21.2 |
| 50 – 64 | 30.2 | 17.0 | 25.9 | 12.0 | 18.5 |
| 65+ | 25.6 | 22.5 | 28.2 | 7.1 | 10.5 |
| Male | 46.3 | 18.7 | 27.2 | 13.1 | 19.2 |
| Female | 53.7 | 10.2 | 19.3 | 6.7 | 14.4 |
| White | 93.2 | 14.9 | 24.1 | 10.2 | 17.3 |
| Non-white | 6.8 | 4.9 | 7.5 | 3.0 | 6.9 |
| Married | 54.4 | 17.6 | 27.2 | 9.3 | 16.3 |
| Not married | 45.6 | 10.1 | 18.0 | 10.2 | 17.0 |
| Income level<sup>b</sup> |  |  |  |  |  |
| Low income | 33.5 | 12.8 | 18.3 | 10.1 | 14.7 |
| Middle income | 31.4 | 12.9 | 23.4 | 8.6 | 16.2 |
| High income | 32.6 | 17.2 | 27.8 | 10.5 | 19.0 |

Note: Estimates are derived from weighted data.
<sup>a</sup> Age groups are based on age reported in April, 2020.
<sup>b</sup> Households earning less than £2,500 a month (net) are classified as low income, those earning £2,500–£4,000 middle income, and those above this threshold as high-income households.

### Regression models

In an adjusted model, the predicted probability of consuming alcohol four or more times per week increased from 14.2 (95% CI[13.5%-14.8%]) to 23 (95% CI[22.2%-23.8%]) between 2017-2019 and April, 2020, a statistically significant increase of 8.8% (95% CI[8%-9.6%]), as outlined in Table 5. Similarly, the prevalence of heavy episodic drinking at least once a week rose significantly from 9.7% (95% CI[9.0%-10.0%]) to 16.6% (95% CI[15.8%-17.4%]) over this period, a significant change of 6.9% (95% CI[6.1%-7.7%]). The increases in both frequent drinking and heavy episodic drinking were statistically significant at the *p* <.001 level for all population subgroups examined (see Table 5) with the exception of non-whites who did not show an increase in drinking four or more times per week.

**Table 5.**
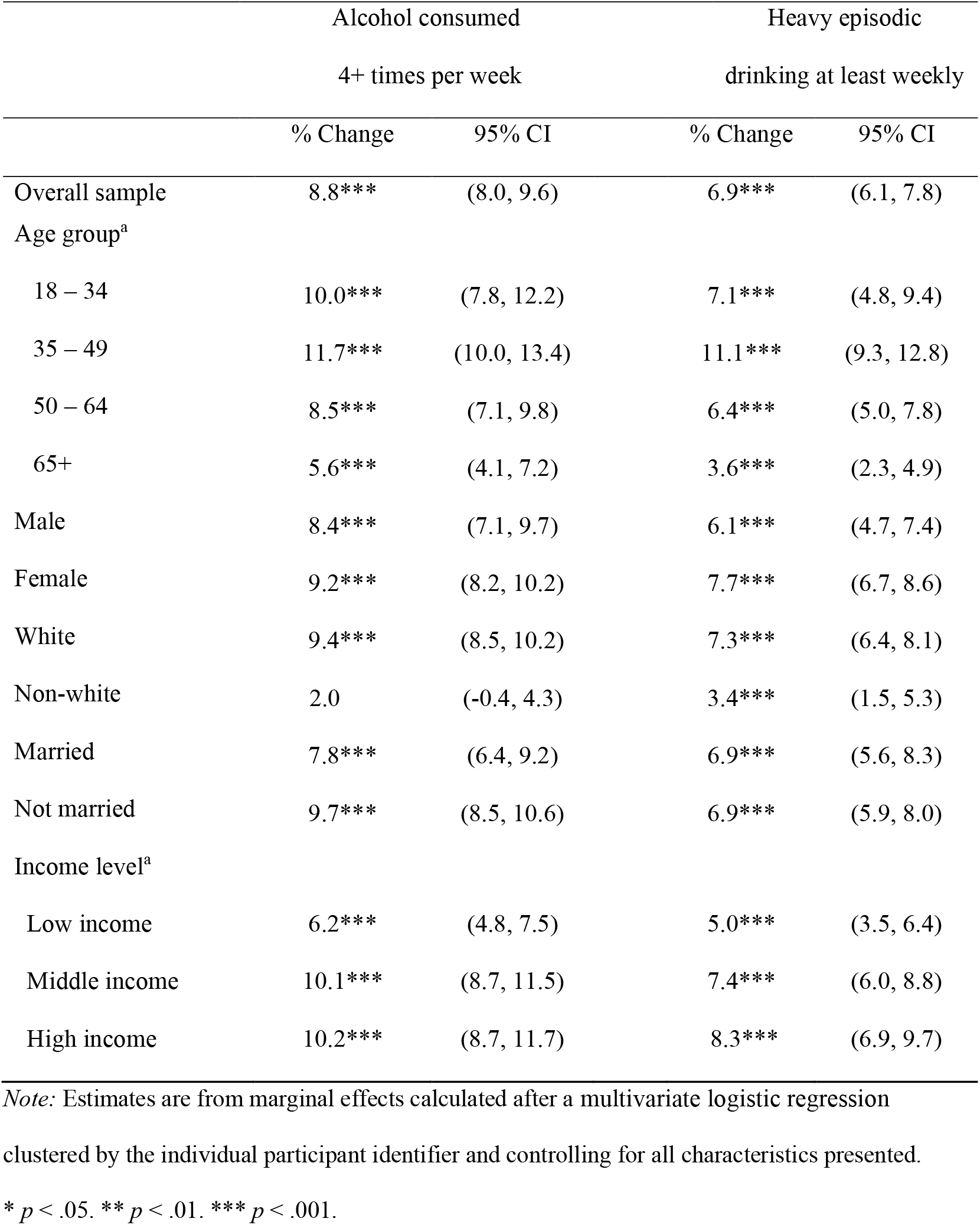
Regression estimates of percentage point changes in problem drinking levels in the UKHLS from 2017-2019 to April, 2020 by population subgroups (N = 12,594; Obs. = 25,188).

The largest increase in frequent drinking (4 times or more a week) was observed among those aged 35-49 years (11.7%, 95% CI[10.0%-13.4%]), followed by those aged 18-34 (10.0%, 95% CI[7.8%-12.2%]). Both groups showed increases that were significantly larger than the increase identified among those aged 65 years and above, as shown in Table 6. Large rises in frequent drinking were also identified among middle income (10.1%, 95% CI[8.7%-11.5%]) and high income (10.2%, 95% CI[8.7%-11.7%]) groups, and these increases were significantly larger than the increase observed in the low income group (see Table 6). The increase in frequent drinking among whites was also significantly larger than the change among non-whites (by 7.4%, 95% CI[4.9%-10.0%]), as shown in Table 6.

**Table 6.** Regression estimates of percentage point changes in problem drinking levels from 2017-2019 to April, 2020 comparing differences between population subgroups (N = 12,594; Obs. = 25,188).

| Variable | Alcohol consumed<br>4+ times per week | Heavy episodic<br>drinking at least weekly |
| --- | --- | --- |
|  | % Change (95% CI) | % Change (95% CI) |
| Age group (comparison is 65+) |  |  |
| 18 – 34 | 4.4** (1.6, 7.1) | 3.5** (0.9, 6.1) |
| 35 – 49 | 6.1*** (3.8, 8.4) | 7.5*** (5.3, 9.6) |
| 50 – 64 | 2.9** (0.8, 4.9) | 2.8** (0.9, 4.6) |
| Male vs. female | -0.8 (-2.4, 0.9) | -1.6 (-3.0, 0.0) |
| White vs. non-white | 7.4*** (4.9, 10.0) | 3.8*** (1.8, 6.0) |
| Married vs. not married | 1.7 (-0.1, 3.6) | 0.0 (-1.8, 1.8) |
| Income level<sup>b</sup> (comparison is low) |  |  |
| Middle vs. low | 3.9*** (1.9, 5.9) | 2.4* (0.4, 4.5) |
| High vs. low | 4.1*** (2.0, 6.1) | 3.3** (1.3, 5.4) |

*Note:* Estimates are from marginal effects calculated after a multivariate logistic regression clustered by the individual participant identifier and controlling for all characteristics presented. Coefficients indicate the difference in the percentage point increase in drinking problems across survey ways between the groups examined.
\* $p < .05$ . \*\* $p < .01$ . \*\*\* $p < .001$ .

The most substantial rise in episodic heavy drinking at least once per week was also among those aged 35-49 years (11.1%, 95% CI[9.3%-12.8%]) and was 7.5% (95% CI[5.3%-9.6%]) greater than the increase among those aged 65 year and over, as shown in Table 6. All age groups experienced increases in episodic heavy drinking that were significantly larger than the increase in the 65+ years age group (see Table 6). Whites also showed a greater increase in episodic heavy drinking compared to non-whites (by 3.8%, 95% CI[1.8%-6.0%]) and those on high incomes showed a larger increase in episodic heavy drinking than those on low incomes (by 3.3%, 95% CI[1.3%-5.4%]).

## DISCUSSION

In the present research we examined changes in problematic drinking among US and UK adults following the development of the COVID-19 crisis. Among US adults, we first examined problem drinking in March 2020 (baseline) when the US death toll of COVID-19 was relatively low (∼5,000) and few states had enacted social lockdown restrictions. We examined the same group of participants across April, when almost all US states had introduced social lockdown restrictions. In this period the proportion of the sample reporting drinking 4 or more times per week increased from 12% to 18%. By May 2020, easing of social lockdown restrictions had occurred across states, but the proportion of the sample reporting drinking 4 or more times per week (17%) remained similar to March levels and was significantly higher compared to baseline. Among UK adults, the baseline assessment was 2017-2019 and 14% of the sample reported drinking four or more drinks per week at this time. Four weeks into social lockdown restrictions in the UK, the prevalence was significantly higher (23%). The UK study we used also included a measure of weekly binge drinking and there was an increase in prevalence from 10% to 17%. Increases in problem drinking were consistently observed across population sub-groups among both UK and US participants. However, there was evidence that more pronounced increases in problem drinking were observed in some sub-groups. In both samples, participants from higher income households showed larger increases in problem drinking. In the UK sample (but not US), there was also evidence that increases were largest among white participants and those ages under 65, with those 35-49 years old group showing the most pronounced increase.

There are a number of plausible mechanisms that may explain population-wide increases in problem drinking. The COVID-19 crisis is thought to have had a considerable burden on population level mental health and this may have resulted in an increase in people using alcohol to cope with stress and negative affect (24, 25). In line with this, a cross-sectional study of alcohol use in COVID-19 social lockdown found that using alcohol to cope was associated with increased drinking in lockdown (26). Likewise, previous research examining how economic crises affect alcohol consumption indicate that rises in psychological distress during times of crisis contribute to increased alcohol use (27).

Furthermore, social lockdown measures have resulted in restrictions in travel, leisure time and physical social engagement, which for many may have resulted in increased boredom. Boredom is thought to have a range of effects on behavior and boredom proneness is linked to higher alcohol consumption (28, 29), which may in part explain why alcohol use has increased alongside the introduction of social lockdown measures.

Across both UK and US samples, higher income participants experienced the largest increases in problem drinking. Associations between socioeconomic status and alcohol use are complex, but higher income tends to be associated with more frequent binge drinking (30, 31). Historical data also suggests that alcohol related harm during times of economic crisis is disproportionately large among the wealthy (32) and more educated (33). During the COVID-19 crisis, it may be the case that existing tendencies towards problem drinking and available material wealth make higher incomes groups more likely to respond to boredom and/or stress by drinking heavily. In a similar vein, increases in problem drinking being larger in white vs. other ethnic groups among UK participants may reflect that abstinence is more common in non-white ethnic groups (34) and such groups would be less likely to use alcohol to cope in times of stress. We also found that among UK (but not US) participants, those under the age of 65 showed the smallest increases in problem drinking. It is plausible that older adults may not be experiencing some of the stressors that younger age groups (e.g. job insecurity due to already being retired, childcare and homeschooling arrangements) will be having to cope with as a result of the COVID-19 crisis.

There are a number of strengths and limitations of the present research. We were able to examine longitudinal changes in problem drinking during the COVID-19 crisis on a person-by-person basis in large nationally representative samples of both UK and US participants. In US participants, the first wave of data collection occurred in March 2020, a period when US states had begun to introduce social lockdown orders and concerns about COVID-19 would have been growing in the US. We conducted sensitivity tests to show increases in frequent drinking were also observed when only participants who were assessed prior to the introduction of lockdown measures in March were examined. Nevertheless, it is plausible that problem drinking had already started to increase at this point, which would result in our analyses underestimating the size of increase in problem drinking associated with the COVID-19 crisis. Likewise, because social lockdown orders were staggered across US states, we cannot attribute overall changes in problem drinking solely to social lockdown orders alone. However, in our UK sample, baseline data was collected prior to the emergence of the COVID-19 crisis and then again after nationwide social lockdown orders.

A limitation of the UK data we used is that baseline data was collected across 2017-2019, whereas follow up data was collected in a single month, though there do not appear to be pronounced seasonality effects on alcohol use in the UK that would explain the sharp rise in problem drinking observed (35). There are also limitations to the measures of alcohol drinking used. Although self-report measures are valid indicators of alcohol consumption, they are prone to bias and error (36). There were also differences in the way that UK participants reported on alcohol consumption at baseline and follow-up. For example, at baseline participants reported using a 12-month time frame, whereas at follow-up the reporting time scale was limited to 4 weeks (to reflect the period of COVID-19 lockdown). It is plausible that reporting error (e.g. underestimation of alcohol consumption) could be larger using a 12-month time frame vs. a 4-week time frame and this may in part contribute to differences in reported alcohol use at baseline vs. follow-up. However, we note that in the US sample the same reporting time frame was used and similar sized increases in problem drinking were observed. As is the case with any longitudinal study there was some level of attrition in both the UK and US samples. In the context of the COVID-19 crisis it is difficult to predict how this may affect estimates of problem drinking. It is conceivable that participants who have developed more substantial alcohol use problems may be more likely to be lost at follow-up and this would underestimate size of change in problem drinking.

## Conclusions

The COVID-19 crisis has been associated with substantial increases in problematic drinking in both US and UK adults.

## Data Availability

Data are available from the dedicated UKHLS and UAS repositiories.

## Acknowledgements

We are grateful to Institute for Social and Economic Research, University of Essex, for their management of the UKHLS data and to the UK Data Archive for making them available. We are also grateful to the University of Southern California’s Center for Economic and Social Research for their management of the UAS data and for making them available. The UKHLS / Understanding Society COVID-19 study is funded by the Economic and Social Research Council and the Health Foundation. However, these organisations bear no responsibility for the analysis or interpretation of the data.

